# Evaluation of the Knowledge of University Students in the Health Area About Sexually Transmitted Infections: A Cross-Sectional Study

**DOI:** 10.1101/2021.02.11.21250842

**Authors:** Carlos Izaias Sartorão Filho, Carlos Izaias Sartorão Neto, Carla Fabiana Souza Guazelli, Camila Rodrigues Avello, Ivana Geraldeli Neiva Rodrigues, Luana Durante Alvarez, Fabiana Vieira Duarte de Souza Reis, Vanessa Patrícia Fagundes, Luiz Takano, Caroline Baldini Prudêncio, Angélica Mércia Pascon Barbosa, Marilza Vieira Cunha Rudge

**Affiliations:** Professor of Medicine at FEMA and student of the Doctoral Program in Toco-gynecology at FMB-UNESP; Medical student at USP-São Paulo; FEMA medical students; Teachers of the FEMA Medicine course; Student of the Doctoral Program in Toco-gynecology at FMB-UNESP; Professor of the Physiotherapy course at UNESP- Marília and the Postgraduate Department in Toco-gynecology at FMB-UNESP; Emeritus Professor of Gynecology and Obstetrics at FMB-UNESP

**Author notes:** Correspondent author: Carlos Izaias Sartorão Filho, Benedito Spinard st, 1440, ZIP 19815-110, Assis – Sao Paulo state – Brazil.

**Keywords:** Health Knowledge Attitudes, Practice, Sexual Behaviors, Sexual Health, Sexually Transmitted diseases, Education, Medical, Undergraduate, Education, Public Health Professional, Health Education

## Abstract

**Introduction:** Sexually transmitted infections (STI) have consequences that negatively affect the population’s quality of life and health. Young adults are susceptible to low access to preventive measures and a greater risk behavior risk. The objective was to evaluate and compare STI’ Knowledge in university students from health courses, a public institution, and a private institution of higher education in Brazil.

**Methods:** a cross-sectional study conducted in 2019 in Brazil at the FEMA School of Medicine in Assis-SP and the University of the State of São Paulo (UNESP) in Marília in the courses of Physiotherapy, Speech Therapy, and Occupational Therapy, in undergraduate students, over 18 years of age, through a self-applied questionnaire translated and adapted to the Brazilian Portuguese from the STD-KQ – Transmitted Disease Knowledge Questionnaire.

**Results:** 462/700 students participated (66.66%) 18-46 years of age (mean 21.46 years, +/− 3.61), 99 men and 363 women. In the private institution, 262/340 (77.06%), and in the public institution, 200/360 (55.55%). The mean age was 22.21 (+/− 4.21) years among men and 21.25 (+/− 3.41) years among women. The mean percentage of the 28 test questions’ correct answers was 52.07%. The average number of correct responses per student was 17.49 out of 28 possible, 62% (+/− 4.97). The distribution between the groups was normal. In the multiple linear regression model for the percentage of correct answers, adjusted for the confounding variables, the intercept was 55.2. The students of the private institution answered 19,655 questions more than the public one (P<.001), and for each semester of graduation, the students answered 1,628 more questions (P<.001). There were no relevant differences regarding the gender and age of the students.

**Conclusion:** there are gaps in STI knowledge among university students in the health area. The students of the first semesters of the courses, and, in particular, students from a public educational institution, had a significantly lower performance of the right in the questionnaire applied. Thus, actions to promote STI knowledge and preventive attitudes are necessary among university students.

## 1. INTRODUCTION

Sexually Transmitted Infections (STI) have a significant impact on sexual and reproductive health worldwide, especially among young people, and are considered a significant public health problem. They produce harmful effects in the short and long term, such as an increased risk of contracting other diseases, infertility, malformations, and deaths (1,2).

The vulnerability of young people is high. However, the perception of risk is significantly low. The low adherence to preventive measures and the early initiation of sexual life make adolescents and young adults more susceptible to these infections (3). Educational actions and counseling adapted to vulnerable populations’ needs are recommended for better preventive and therapeutic STI control (4).

In this sense, assessing young people’s knowledge about STI allows identifying knowledge gaps that may predispose them to greater vulnerability since experience is the basis for people’s attitudes and practices. This assessment provides relevant support for the formulation and implementation of actions that encourage and support conscious preventive behaviors. (1–3)

## 2. OBJECTIVES

### 2.1 Primary objective

To evaluate the Knowledge of STD students in the health area about STI.

### 2.2 Secondary objectives

To investigate preferential distributions according to gender, age, and the semester in which the students are in their courses.

To compare results between a public education institution and a private institution.

## 3. HYPOTHESES

There are knowledge gaps in the STI theme among higher education courses in the health area.

## 4. METHODS

This is a cross-sectional study carried out on undergraduate students of the medical course at FEMA - Educational Foundation of the Municipality of Assis, in the State of São Paulo, a private educational institution, and on undergraduate students in the courses of Physiotherapy, Occupational Therapy, and Speech Therapy at UNESP-Universidade Estadual Paulista, Campus de Marília, State of São Paulo, a public educational institution. The Research Ethics Committee (Certificate number: 13034619.0.0000.8547) previously approved the study. We carried recruitment and data collection out in April and May of the year 2019 through a self-administered questionnaire to all students of the institutions who consented to participate. A free and informed consent form with the clarifications was obtained before applying the questionnaire. The questionnaire was translated and culturally adapted into Brazilian Portuguese from the original English version (5) of the STD-KQ - Sexually Transmitted Disease Knowledge Questionnaire scale (6). An item about lambskin condom was removed from the instrument since this material is not found in Brazil. Also, to contemplate the Brazilian epidemiological scenario, the researchers added two questions about syphilis. The Brazilian version comprises 28 statements about seven STI: syphilis, gonorrhea, chlamydia infection, genital herpes, HPV, HIV / AIDS, and viral hepatitis. It categorizes the questionnaire responses as true, false, and I don’t know. A study of the psychometric evaluation of the Brazilian version of the STD-KQ showed internal consistency (composite reliability = 0.97; Cronbach’s alpha = 0.83) and temporal stability (Pearson’s correlation = 0.86; kappa = 0.16) for one short period. In a single day, the taking part students received the classroom questionnaire in a brown envelope, without identification. After filling anonymously, the envelopes were returned, sealed, and collected for later analysis and tabulation. The students’ knowledge was obtained by adding the correct answers, with a minimum of 0 and a maximum of 28 for each participant.

### 4.1 Inclusion and exclusion criteria

#### Inclusion criteria

students regularly enrolled in the medical course of the Educational Foundation of the Municipality of Assis and the Physiotherapy, Occupational Therapy, and Speech Therapy courses at Universidade Estadual Paulista; age equal to or over 18 years old; non-participants as author or co-author of the study.

#### Exclusion criteria

illegible questionnaires, erasures, not filled in correctly, or returned outside the sealed envelope.

### 4.2 Sample size

A minimum representative sample size of 162 adolescents and 269 adults was calculated from the equation for a nonprobabilistic sample for an infinite population-based on a proportion of 12% of 10-19-year-old and 50% of adults in São Paulo state. A confidence interval of 95% and an error size of 5% were used.

### 4.3 Data analysis methodology

We analyzed the data using SPSS software version 24 (Statistical Package for the Social Sciences-IBM). We presented descriptive results in tabular form. The level of statistical significance was determined as p <.05. It described the Age variable in years and calculated the Average Age and Standard Deviation. It described variable Educational Institution as private (FEMA) and public (UNESP), and we divided the graduation stages into semesters studied. We have described the gender in Male and Female. The outcome comprised 28 multiple-choice questions, containing three answers: yes, no, and I don’t know. Only one answer in each question was considered correct. Each valid question received a score of 1 point. Incorrect answers or “I don’t know” or unanswered received a score of zero points. The average sum of the 28 questions was calculated as a percentage of correct answers compared with the study variables, age, sex, institution, and the semester of the course.

Statistical analysis used parametric and non-parametric tests for compare groups, and the multiple linear regression model for the percentage of right answers, adjusting potential confounders. Through this analysis, for the variables public institution, semester, age, and male gender, the beta coefficient (β), the power of significance (p), and the 95% confidence interval (95% CI) were calculated, and the variance inflation factors (VIF).

## 5. RESULTS

The total number of participants was 464 students, 2 of whom it excluded from the survey for inadequate questionnaire filling. The 462 students’ age ranged between 18-46 years (mean age 21.46 years, +/− 3.61), with 99 men and 363 women. The study’s overall participation rate was 54.28% (360/700). At FEMA, 77.06% (262/340), and at UNESP, 55.55% (200/360). In stratification by sex, the mean age was 22.21 (+/− 4.21) years for men and 21.25 (+/− 3.41) years for women. There was no refusal to take part in the study. The enrolled students who did not participate were absent from academic activities because we administered the questionnaire at each institution.

At FEMA, of the 262 students who took part, 182 were women and 80 men. FEMA students’ average age was 21.55 years (+/− 3.39); the average percentage of correct answers for the 28 test questions was 70.39%. At UNESP, 200 students took part, 181 women and 19 men. The participants’ average age was 21.32 years (+/− 3.89). The average percentage of correct answers for the 28 test questions was 52.07%.

The global average of correct answers per student was 17.49 out of 28 possible, or 62%, and the standard deviation was 4.97. The overall average of correct answers per question was 288.57 out of 462 possible, making up 62%, and the standard deviation of 88.30.

Graph 1 shows the total number of correct questions and the percentage of correct answers for each question. Table 1 shows the number of students taking part in each stage/semester of the courses. Table 2 shows the multiple linear regression model for the percentage of correct answers, adjusted by the confounding variables.

**Table 1.**
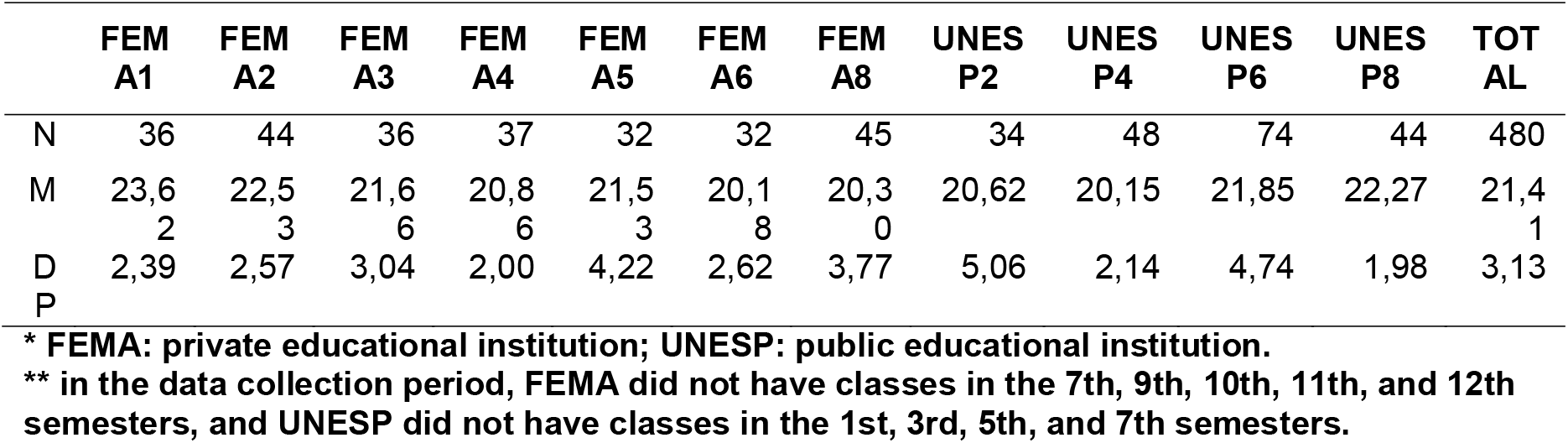
**The number of students per institution and semester (N) means age (M) and Standard Deviation (SD).**

**Table 2.** **Multiple linear regression model for the percentage of the correct answer, adjusted by the confounders.**

| Variável | $\beta$ | p | 95%CI | | VIF |
| --- | --- | --- | --- | --- | --- |
| Intercept | 55,204 | ,000 | 46,950 | 63,458 |  |
| Public institution (Ref: Private) | -19,655 | ,000 | -22,563 | -16,747 | 1,146 |
| Semester | 1,628 | ,000 | ,990 | 2,266 | 1,146 |
| Age | ,364 | ,067 | -,025 | ,752 | 1,087 |
| Male | 1,850 | ,287 | -1,561 | 5,261 | 1,082 |

**Graph 1.**
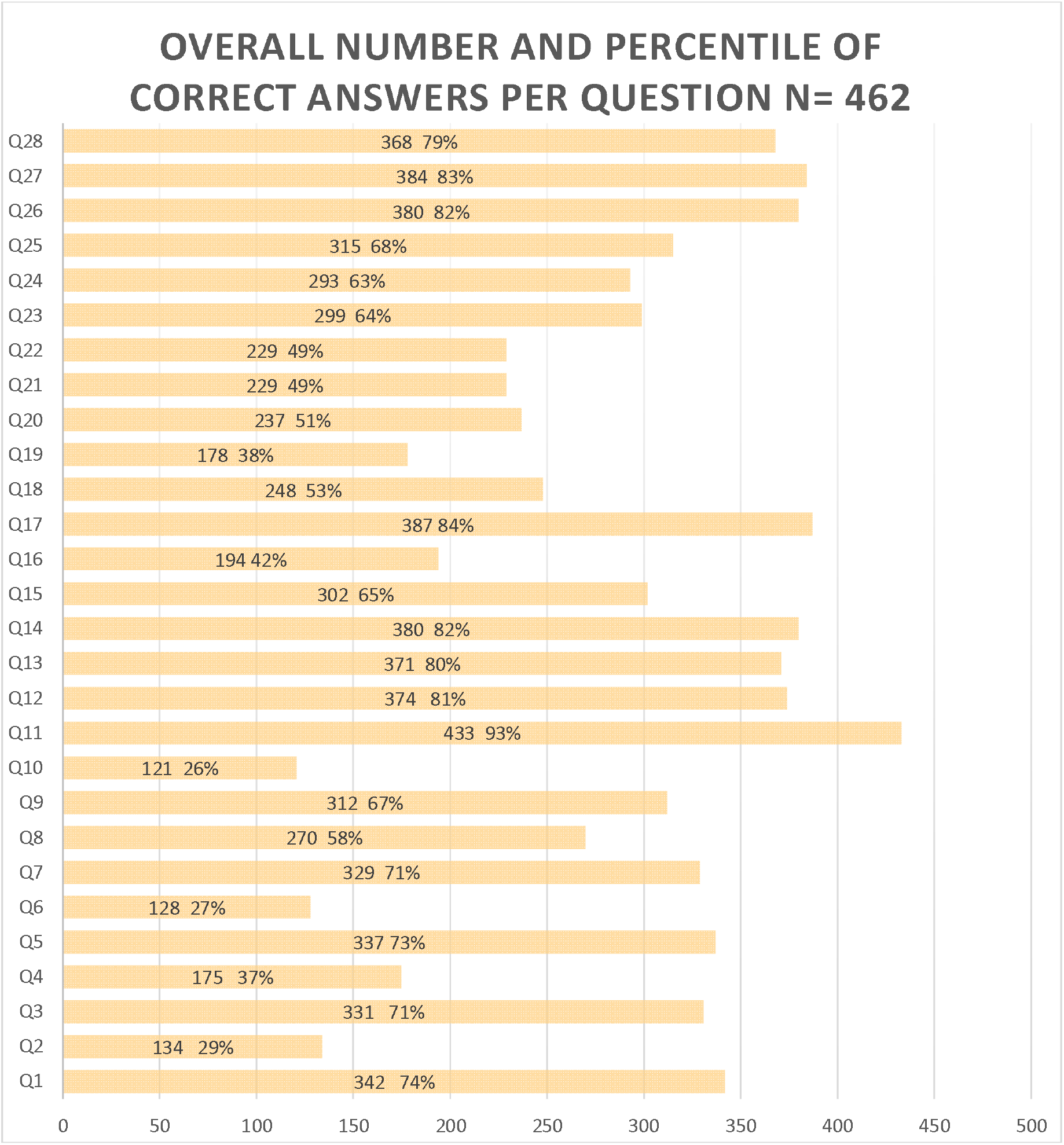
The total number of correct questions and the percentage of correct answers to each question. The total mean number of correct answers was 64.46% (+/− 17.76%).

The Post-hoc Power analysis was 98.1%, based on the percentage of correct answers between the institutions and the type 1 error percentage of 5%.

## 6. DISCUSSION

The study results, demonstrated by a global average of correct answers per student of 62% (+/− 4.97), denoted gaps in knowledge among university students from public and private institutions in the health area regarding the theme IST.

It is noteworthy that many questions had a low rate of correct answers. In the percentage analysis of correct answers for each question, we had eight questions with a global index of correct answers below 50%. Thus, the possibility of the studied group being exposed to risk practices and attitudes concerning STI’ contagion and transmission.

When analyzing the percentage of correct answers applied, using a multiple linear regression model, adjusting the confounding variables, it is observed that the intercept was 55.204 (95% CI 46.950-63.458), statistically relevant (p <.001). Concerning students from private institutions, the Beta coefficient was −19.655 (95% CI −22.563 − 16.747), a significantly relevant value (p <.001). Students from the public institution answered 19.65% fewer questions on average than those from the private institution. As for the stage/semester of the course, the Beta coefficient was 1.628 (95% CI, 990 - 2.266), a significant value (p <.001). Each semester attended, it is estimated an increase of 1.628 percentage points of correctness, which is significant. As for the age and sex variables, there were no significant changes.

There were only two exclusions from the study because of the lack of filling in the questionnaire, delivered with no answers. No student refused to participate in the survey during recruitment at the institutions. The acceptance rate for participating in recruitment and appliance was 100% at both institutions.

The study recruited university students from two medium-sized cities, Assis and Marília, in the interior of the State of São Paulo, Brazil, covering courses in the areas of Medicine, Speech Therapy, Physiotherapy, and Occupational Therapy, in public and private institutions. It is also noteworthy that we applied the questionnaire only to university students in the health area, so it was estimated that they achieved more knowledge about the STI theme. Thus, it is believed that the results found can be generalized and extrapolated to the population of university students in the country, regardless of the discipline or area of Knowledge studied.

Study limitations: the literature not validated the Portuguese language questionnaire. Therefore, different interpretations of the statement and the answers to each question may generate inaccuracies. Another rule is that the questionnaire was applied to students of the private institution’s medical course. In contrast, the questionnaire was used in the public institution to students in Physiotherapy, Occupational Therapy, and Speech Therapy courses. There are differences in these courses’ curriculum, causing potential bias, depending on subjects that may have previously addressed the IST theme or taking part in classes and institutions. Despite this, we consider that the questionnaire assesses students’ prior knowledge about STI. This evaluation is essential for recognizing potential needs and developing strategies to understand infections better.

Samkange-Zeeb et al. (7) published a systematic review in 2011 on knowledge and awareness among European adolescent students regarding Sexually Transmitted Diseases. Between 1990 and 2010, the authors selected 15 studies, all of them transversal, with students between 13 and 20 years of age. Awareness was high about HIV / AIDS (over 90%) and low for HPV (5-66%). Overall, meta-analysis studies reported low awareness and Knowledge about STI among adolescents.

A study carried out in Brazil by Fontes et al. (3) with 1208 young people between 18 and 29 years old from 15 Brazilian states assessed determinants of knowledge, attitudes, and practices about STI. The primary variable was a scale with 35 questions on the topics. Regarding experience, 40% answered that they did not consider condom use as a restorative measure to prevent STI or pregnancy. Regarding attitude, 40% believed that there is no longer a need to use condoms in a stable relationship. Less than 10% of respondents, sexually active, had sought a health service in the past 12 months to consult prevention. In conclusion, young Brazilian adults have shown insufficient knowledge and ineffective STI attitudes.

Research developed by Castro et al. (8) by the University of Campinas aimed to quantify and generate self-perception about STI and to test students’ interest in creating a discipline on the subject. The data collection instrument was the questionnaire sent electronically to undergraduate students at the end of 2011 and newly enrolled students in 2012. 99% of sexually active students used the condom, but less than 20% used it correctly. About 80% were unaware that the condom does not protect outside the barrier area; they intended to read more about IST and learned something about it. Almost half thought that discipline should be offered to all graduates.

Almeida et al. in 2017 in Brazil (9) investigated the knowledge of high school teenagers in public schools. They found the need for preventive educational actions because of the low level of information received in four thematic categories: sexuality and sexuality education, understanding of risk behaviors, Knowledge about STI, and knowledge and practices of STI prevention.

A study carried out among university students in Health in Geneva, Switzerland, identified that students of both sexes have low Knowledge about HPV infection, its forms of contagion, and methods of preventing the disease (10).

Visalli et al. (11) evaluated students’ knowledge and risk behavior about STI in Italian schools and universities. The results showed an extreme variability of expertise. Multiple linear regression showed that knowledge about sexual health was associated with age and sexual orientation. The results showed a low or inadequate understanding of students about STI, findings that supposedly would put them in situations of vulnerability.

In a cross-sectional study in 2018, Provenzano et al. (12) assessed sexual and reproductive risk behavior among Italian university students using an online questionnaire. Among the 539 students who took part, the average age was 22.65 (+/− 2.95) years. They reported that it associated the risk of having casual unprotected sex with the following circumstances: homosexuality and bisexuality, male gender, age at sexarche under 17, and a history of having STI previously. They concluded in their analysis that it is necessary to implement sex education programs and that university students decrease precautions about STI.

In 2018, Santangelo et al. (13) assessed STI and sexual risk behavior among Italian health students. Then, 1022 respondents were analyzed, most women and aged between 18 and 22 years. In the multivariate analysis, men were at greater risk of not knowing about the HPV vaccine; the age group between 18 and 22 years had a higher chance of not being sufficiently informed about how to avoid contagion, of never having had serological tests, of not knowing the HPV vaccine, and not knowing that they can use the HPV vaccine in men. The authors concluded that it is necessary to implement programs to increase knowledge about STI and promote health, including young adults who have not entered universities.

## 7. CONCLUSIONS

There are knowledge gaps in adolescents and young adult university students in the health area about STI. Besides, students from the first semesters of the courses and, in particular, students from a public educational institution, had an inferior performance in the applied questionnaire. As for age and gender, there were no significant changes. These findings are essential for schools and graduation institutions to develop educational and awareness strategies to mitigate STI risks among students. Thus, the discussion on addressing the theme of Sexually Transmitted Infections in the curriculum, especially in the first stages of the courses, is mandatory.

## Data Availability

Data availability statement: we declare that all data referred to in the manuscript can be available under request.

https://www.dropbox.com/s/6htpbib2d2h0kuw/ISTpreprint.docx?dl=0

## 8. FINANCING

none.

## REFERENCES

1. Unemo M, Bradshaw CS, Hocking JS, de Vries HJC, Francis SC, Mabey D, et al. Sexually transmitted infections: challenges ahead. Lancet Infect Dis [Internet]. 2017 Aug;17(8):e235–79. Available from: https://linkinghub.elsevier.com/retrieve/pii/S1473309917303109

2. McCormack D, Koons K. Sexually Transmitted Infections. Emerg Med Clin North Am [Internet]. 2019 Nov;37(4):725–38. Available from: https://linkinghub.elsevier.com/retrieve/pii/S0733862719300732

3. Fontes MB, Crivelaro RC, Scartezini AM, Lima DD, Garcia ADA, Fujioka RT. Fatores determinantes de conhecimentos, atitudes e práticas em DST/Aids e hepatites virais, entre jovens de 18 a 29 anos, no Brasil. Cien Saude Colet [Internet]. 2017 Apr;22(4):1343–52. Available from: http://www.scielo.br/scielo.php?script=sci_arttext&pid=S1413-81232017002401343&lng=pt&tlng=pt

4. Kalamar AM, Bayer AM, Hindin MJ. Interventions to Prevent Sexually Transmitted Infections, Including HIV, Among Young People in Low- and Middle-Income Countries: A Systematic Review of the Published and Gray Literature. J Adolesc Heal [Internet]. 2016 Sep;59(3):S22– 31. Available from: http://www.jahonline.org/article/S1054-139X(16)30101-X/pdf

5. Jaworski BC, Carey MP. Development and psychometric evaluation of a self-administered questionnaire to measure knowledge of sexually transmitted diseases. AIDS Behav. 2007;

6. Teixeira LO, Figueiredo VLM de, Gonçalves CV, Mendoza-Sassi RA, Teixeira LO, Figueiredo VLM de, et al. Avaliação Psicométrica da versão brasileira do “Questionário sobre Conhecimento de Doenças Sexualmente Transmissíveis.” Cien Saude Colet [Internet]. 2019 Sep [cited 2019 Dec 15];24(9):3469–82. Available from: http://www.scielo.br/scielo.php?script=sci_arttext&pid=S1413-81232019000903469&tlng=pt

7. Samkange-Zeeb FN, Spallek L, Zeeb H. Awareness and Knowledge of sexually transmitted diseases (STDs) among school-going adolescents in Europe: a systematic review of published literature. BMC Public Health [Internet]. 2011 Dec 25 [cited 2020 Dec 20];11(1):727. Available from: http://bmcpublichealth.biomedcentral.com/articles/10.1186/1471-2458-11-727

8. Castro EL de, Caldas TA de, Morcillo AM, Pereira EM de A, Velho PENF. O conhecimento e o ensino sobre doenças sexualmente transmissíveis entre universitários. Cien Saude Colet [Internet]. 2016 Jun;21(6):1975–84. Available from: http://www.scielo.br/scielo.php?script=sci_arttext&pid=S1413-81232016000601975&lng=pt&tlng=pt

9. Almeida RAAS, Corrêa R da GCF, Rolim ILTP, Hora JM da, Linard AG, Coutinho NPS, et al. knowledge of adolescents regarding sexually transmitted infections and pregnancy. Rev Bras Enferm [Internet]. 2017 Oct [cited 2020 Oct 4];70(5):1033–9. Available from: http://dx.doi.org/10.1590/0034-7167-2016-0531

10. Jeannot, Viviano, Follonier, Kaech, Oberhauser, Mpinga, et al. Human Papillomavirus Infection and Vaccination: Knowledge, Attitude and Perception among Undergraduate Men and Women Healthcare University Students in Switzerland. Vaccines [Internet]. 2019 Sep 26 [cited 2020 Oct 4];7(4):130. Available from: https://www.mdpi.com/2076-393X/7/4/130

11. Visalli G, Cosenza B, Mazzù F, Bertuccio MP, Spataro P, Pellicanò GF, et al. knowledge of sexually transmitted infections and risky behaviours: a survey among high school and university students. J Prev Med Hyg [Internet]. 2019 Jun [cited 2020 Oct 4];60(2):E84–92. Available from: https://doi.org/10.15167/2421-4248/jpmh2019.60.2.1079

12. Provenzano S, Santangelo OE, Alagna E, Giordano D, Firenze A. Sexual and reproductive health risk behaviours among Palermo university students: results from an online survey. Clin Ter. 2018;169(5):242–8.

13. Santangelo OE, Provenzano S, Firenze A. Knowledge of sexually transmitted infections and sex-at-risk among Italian students of health professions. Data from a one-month survey. Ann Ist Super Sanità. 2018;54(1):40–8.

